# Managing COVID-19 in an Australian designated isolation facility: Implications for current and future healthcare crises

**DOI:** 10.1101/2022.05.05.22274702

**Authors:** Helen M Achat, Rakhi Mittal, Joanne M Stubbs, Nicky Gilroy, Suzanne K Schindeler, Ramon Z Shaban, Thomas Solano

## Abstract

Health care workers’ (HCWs) lived experiences and perceptions of the pandemic can prove to be a valuable resource in the face of a seemingly persistent Novel coronavirus disease 2019 (COVID-19) – to inform ongoing efforts, as well as identify components essential to a crisis preparedness plan and the issues pertinent to supporting relevant, immediate change. We employed a phenomenological approach and, using purposive sampling, conducted 39 semi-structured interviews with senior healthcare professionals who were employed at a designated COVID-19 facility in New South Wales (NSW), Australia during the height of the pandemic in 2020. Participants comprised administrators, heads of department and senior clinicians. We obtained these HCWs’ (i) perspectives of their lived experience on what was done well and what could have been done differently and (ii) recommendations on actions for current and future crisis response. Four themes encapsulated respondents’ insights that should inform our capacity to meet current needs, direct meaningful and in situ change, and prepare us for future crises. Respondents’ observations and recommendations are informative for decision-makers tasked with mobilising an efficacious approach to the next health crisis and, in the interim, would aid the governance of a more robust workforce to effect high quality patient care in a safe environment.

## Introduction

Novel coronavirus disease 2019 (COVID-19) provided irrefutable evidence that the world is unprepared for outbreaks. Actionable plans were noticeably wanting in March 2020 when the World Health Organization (WHO) declared a global pandemic. COVID-19’s rapid spread has challenged entire health care systems globally to meet the requirements of patient care while simultaneously protecting their health care workers (HCWs). As with previous outbreaks such as Severe acute respiratory syndrome (SARS) and Middle East respiratory syndrome coronavirus (MERS-CoV), HCWs have been facing extraordinary demands related to high risks of infection, stigmatisation, understaffing and uncertainties about the virus.[1, 2] Acknowledgment that their health and safety is crucial – for workers themselves, the continuing and safe care of their patients, and control of outbreaks,[3] – has not manifested into concrete measures that minimise HCWs’ risk of mental health issues,[4-6] infection and death from COVID-19.[7]

Nearing 24 months since WHO’s declaration, countries worldwide continue to struggle with adequate reliable health care in the face of the pandemic.[8, 9] Pandemic-distinct challenges, known but some elusive to overcoming, include matters related to adequate supply and effective use of personal protective equipment (PPE),[10, 11] the safety and well-being of an appropriately skilled health workforce,[3, 12, 13] workplace structures, practices and policies that shape the experiences and capabilities of HCWs[14, 15] and an overarching adaptable decision making and management approach.[16, 17]

Australia, with a population of around 25 million, experienced low infection and death rates compared to most OECD countries prior to the Omicron COVID-19 variant, which resulted in exponential rise in new cases and a notable increase in deaths.[18] Its relative success till late 2021, attributed in part to its structural advantage as an island nation able to close its boarders[19] and strong collaboration among experts, by no means spared HCWs and the public from the aforementioned challenges in early 2020. As all countries come to terms with a seemingly inevitable ‘COVID normal’ state of operations, the lived experience and perceptions of HCWs who held senior clinical and administrative positions during the pandemic can prove to be a valuable resource. Frontline responders’ unique insights into health care delivery can enable discernment of aspects of health care and its coordination that have been efficiently addressed, areas that prove to be ongoing challenges, as well as reasons for the situations encountered. Explicating the lessons learnt should galvanise decision makers to meet current needs and identify issues relevant to a preparedness plan for future crises.

The aim of this study is to document, through the garnering of lived experiences and perceptions, Australian HCWs’ considerations on what they view as intrinsic to effective health care measures for a successful pandemic response, and in so doing identify those aspects of health care that should be sustained or adapted going forward. This study focuses on HCWs, non-clinical and clinical, from a designated COVID-19 facility in New South Wales (NSW), Australia.

## Materials and Methods

### Study design

This qualitative study was conducted using a phenomenological approach through individual semi-structured in-depth interviews. Addressing the study questions “What could have been done, or done differently to better respond to the pandemic?” and “What would you like to see maintained as we move towards a ‘COVID-19 normal’ phase?”, researchers aimed to describe staff’s perspective of changes in their lived experience at the height of the pandemic in 2020 and their recommendations on actions that could facilitate their workplace’s response to COVID-19 and preparedness for future health crises.

### Procedure

In-depth, semi-structured individual interviews were conducted in person or via online video (Zoom), according to the participant’s choice. The interviewers were three of the co-authors all of whom were female and qualified and experienced public health professions. Participants were asked a preliminary question about their roles prior to and subsequently during the height of the pandemic, which enabled confirmation of their involvement in the hospital’s pandemic response. The interviewer then loosely followed a script that supported in-depth discussion of respondents’ lived experiences and perceptions on the subjects of staff stress and anxiety, changes in workplace policies and procedures, communication methods, challenges, areas for improvement and recommendations for healthcare response and preparedness. The interviewer used probing questions to gain more in-depth responses and aid clarification. All interviews were recorded with the interviewees’ consent.

### Participants and setting

All participants worked at a large tertiary teaching hospital that is a designated COVID-19 management facility in NSW, Australia, which received confirmed COVID-19 patients from late January 2020. We employed purposive and snowball sampling (see Table 1 for a description of the professional groups), which was supported by the Director of Infectious Diseases and Prevention and the Clinical Nurse Consultant and lead of Infection Control at the hospital. They each compiled a list of potential key informants comprising hospital staff employed during the height of the COVID-19 pandemic. Each key informant was sent a letter of introduction with details of the study and the contact details of the Principal Investigator. Subsequent communication by telephone and email confirmed consent to participate in a semi-structured interview with an expected duration of approximately 45 minutes and an interview appointment.

**Table 1.**
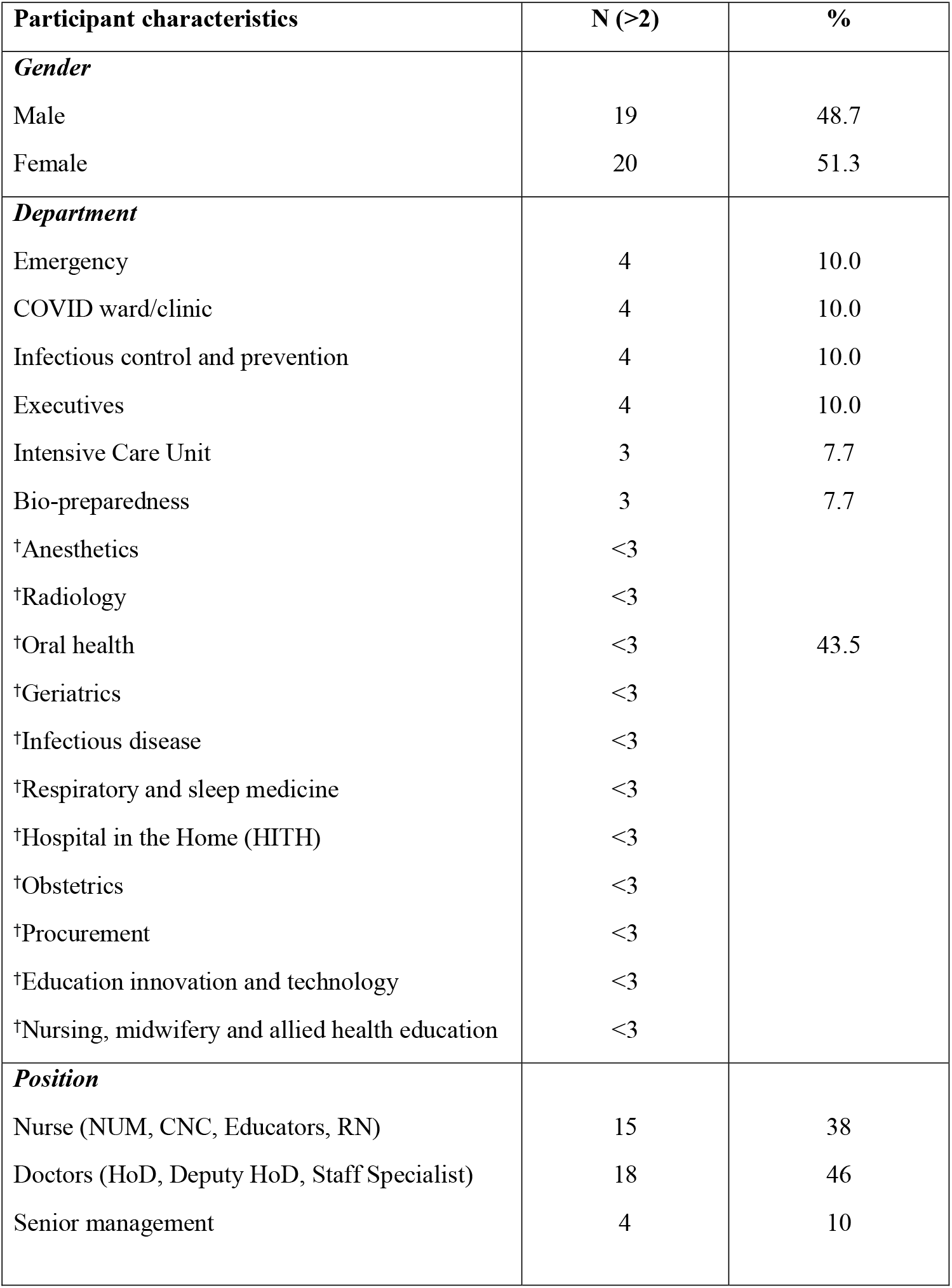
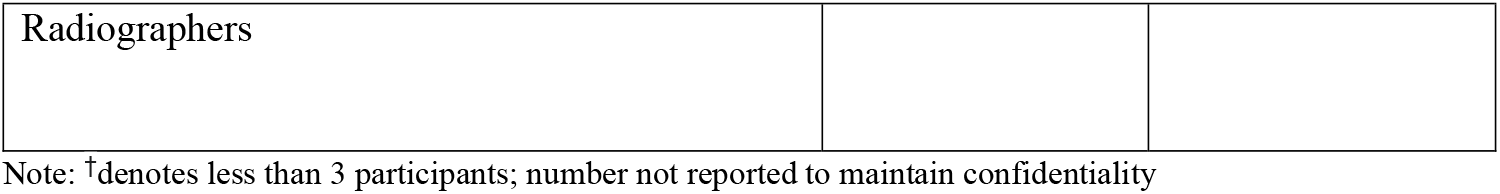
Characteristics of participants (N=39)

### Data analysis

Interview audio-recordings were imported into a commercial program that produced a rough transcription. The interviewer cleaned the initial transcript by assigning speakers to the script and correcting text and punctuation.

Data were analysed using Colaizzi’s distinctive seven step process [20] that provides a rigorous analysis, with each step staying close to the data, which includes familiarisation, identifying significant statements, formulating meanings, clustering themes, developing an exhaustive description, producing the fundamental structure and seeking verification of the fundamental structure. Researchers engaged in ongoing analyses. Transcripts were analysed by at least two researchers using a standard template, before discussions with the wider research team about emerging themes to inform the focus of subsequent interviews.

Following the transcription of the first few interviews, a review of the completed templates by research team members confirmed persistent themes. The team was responsive and open to what was in the data and allowed it to guide an iterative approach to analysis. Researchers analysed the transcripts independently by bracketing data on preconceived ideas and strictly following the Colaizzi method described above. Early analysis allowed the initial development of a coding framework that underwent ongoing development as transcripts were re-read and reviewed. Researchers reviewed emerging findings at regular team meetings until data saturation occurred and used consensus to resolve disagreements.

A key to reading the quotes is provided below:

1. [] indicates the researcher has added the narrative to make the context and/or meaning clearer, or replaced some identifiable text with de-identifiable text.
2. … means that words, phrases or sentences of the interview have been deleted to make the context and/or meaning clearer.
3. () after each original participant quote indicates that particular participant’s numerical pseudonym.

### Rigor/trustworthiness

The criteria of credibility, dependability and conformability [21] were used to confirm the rigor of the findings. Following the transcription of the first few interviews, a peer review process was used by the research team members to confirm persistent themes. We adhered to all assumptions and strategies of the qualitatively driven designs. We were responsive and open to what was in the data and allowed the data to guide our iterative approach to analysis. We reviewed emerging findings during daily team meetings to ensure data saturation and consensus among study team members. This was done to ensure the credibility of research.

### Ethical considerations

This study was approved by Western Sydney Local Health District Ethics Committee (2020/ETH01674). All participants received information about the study, and a consent form, which they were asked to read, sign and return. They were made aware that they could refuse to answer any question and could terminate the interview at any time. Our study adhered to standard ethical processes for qualitative research to ensure the anonymity of participants and confidentiality of the data.

## Results

### A description of participants and interviews

Thirty-nine interviews were conducted via Zoom (N=35) and face-to-face (N=4). All participants were employed in roles with a primary focus on the provision of health care in the height of the pandemic. We obtained over-representation from the emergency department (ED), intensive care unit (ICU), respiratory medicine and infectious diseases (ID) departments including the COVID clinic and ward, which were deemed high exposure environments. Twenty were in newly created roles working either on the COVID-19 ward or in another capacity (Table 1). Interviews had a duration of 30 to 104 minutes, averaging 60 minutes and were conducted from November 2020 to February 2021.

### Key considerations for an effective response

The unknown characteristics of the virus, initially amplified by a plethora of information from local and international sources, were a cause of heightened anxiety and stress. HCWs were acutely affected by a number of factors, among them the potential shortage of and education about PPE use, a tremendous COVID-19 related workload, leave cancellations, and moral dilemmas around the provision of less-than-optimal care and the social isolation of patients. The initial absence of guidelines and anticipated directives from the Ministry of Health stymied the process of informed decision-making. Participants had a range of experiences of and considerations about measures undertaken or lacking to address safe health care delivery.

Four themes encapsulated respondents’ lived experiences and reflections on what was done well, what could have been done differently, and considerations for ongoing and future provision of health care (Figure 1): minimising the spread of disease at all times, maintaining a sense of collegiality and promoting informed decision-making, maintaining a plan for future crises and promoting corporate and clinical agility. We have mentioned selected quotations throughout the Results and have presented detailed quotations in Table 2.

**Table 2.** Detailed quotes categorised by themes.

| Theme | Illustrative quotes |
| --- | --- |
| **Theme 1. Minimise the spread of disease at all times** |  |
| a. Centralised and managed product procurement including PPE. | *If we had a CELP [centralised equipment loan pool], or a centralised thing, if someone was in charge of maintaining that, we wouldn't have expired stock. (P02)* |
| b. Under the WH&S umbrella, enhance/maintain necessary infection control measures and training | *I don't think I've ever seen better hand hygiene in the hospital, wiping surfaces down, making sure that we're always keeping things clean and tidy, respiratory etiquette, coughing into the elbow, it's a really nice thing ... (P27)*<br>*We made some substantial changes to clinical practice around here over that time that, going forward, you'd like to see maintained ... we have all these protocols and procedures now... In the event of something else like this, we have to be careful we don't lose them, because they're all done and they all work ... a greater awareness by staff on PPE use, and what's most effective with that. I think that's been really good. (P11)* |
| c. Provide consistent staff with suitable training | *They could have given me permanent staff in both areas, instead of this constant fight for staff and constant education and constant training. (P14)*<br>*One of the hardest things from the very start, I felt, from a frontline staff health perspective, is ... they had to staff it [the COVID ward] with in-house pool staff, or agency staff ... And we've obviously learned as time has gone on that was not a good thing, because there was hardly any continuity of staff on each shift. There was no person put in charge of that ward [initially] ... They desperately needed somebody on that ward to pull them together ... (P36)* |
| d. Continue limits on visitor numbers | *The change in visiting policy was [something] everyone struggled with very much, even me. ... When the visiting changed, the visiting policy was absolutely zero visitors. Especially when you had dying COVID patients – that was really challenging. A lot of staff were quite traumatized by that ... We had, how many deaths? ... In one family the wife was allowed in for half an hour, and in the other one [the patient] died alone. So that is ... very hard for the nurses to accept. Well, everyone really ... this is really against all our practices, our morals, our values. That was hard to swallow, but I guess it's the greater good, for the greater amount of people and it's to protect everyone. But ... when you're so entrenched in your morals, values and how death should look, that was very hard to overcome. (P22)* |
|  | *Keeping the visitor [numbers] small, because ... unless there's a few special circumstances ... generally, most people only need one person with them, or two max, and probably actually need to have a rest because they are sick. So just keeping numbers low, we'll probably continue that. (P04)* |
| e. Provide fit for purpose infrastructure to reduce transmission in hospital (e.g. hot zones, separate COVID clinic and ward, negative pressure rooms) | *I think the built physical environment is very critical. Just due to the nature of our work and the potential for airborne transmission. (P13)*<br>*You need to understand better how to manage these airborne transmissible diseases in a hospital setting, and do it effectively all the time, not just because the pandemic has come around ... it just makes no sense to me anymore to be 'cohorting' infectious patients together. (P11)*<br>*So, we had to then reconfigure the emergency department and build a negative flow room. And then, how do you treat patients in these rooms? How do you communicate with them? How do you communicate with the staff? And how do you respond, doing your straightforward work, such as giving morphine, for example. That was a huge, huge task. (P34)* |
| f. Maintain processes and procedures that support infection control and WH&S | *Some good stuff came out of it [the pandemic]. We've learnt how to make special circuits for NIV for the infection-prone ... NIV circuits that had expiratory valves on them, so that you could filter the air that was coming out. And that was quite big, because you have to change the whole circuit setup. There was a lot of work done by a few people to figure out how to have these safer circuits that we could use if we needed to ... Stopping use of nebulizers, better controlled use of oxygen or high-flow oxygen, those were all important things. (P11)* |
| **Theme 2. Maintain a sense of collegiality and informed decision-making** |  |
| a. Preserve teamwork and working out of silos as displayed during pandemic | *It was a great response in the end and I think everybody was happy to knuckle down and put in what was going to be required, recognising that it might involve a lot of extra work and time to deal with this problem if it gets out of control ... a sense of 'well, that's what our job would be'. (P11)*<br>*I think in a pandemic ... as a hospital you come together, everyone's got one agenda ... so you pulled together as a team, and that really occurred; everyone put aside their [own] agenda. (P24)*<br>*[We] have an intensive care department and emergency department and anaesthetics department who all talk to each other now. It's actually changed communication. (P21)* |
| b. Acknowledge the contributions of all teams of healthcare workers | *For us, it was important to acknowledge staff for their work and effort, and it was an eye-opener how some people responded, and how much value they added. (P31)* |
|  | *Now we're coming down to a normal period. I think it's also a time that staff can sort of receive that word of appreciation, that gesture of appreciation. (P09)* |
| c. Activate one source of information to reduce overload and excessive channels of information | *There's communication, and how you get communication to every tier of your organisation in a way that's consistent. The messaging has to be consistent ... that is very important. We have what we hear on the TV, what we hear at a national level, what we hear at a jurisdictional level, what we hear at the hospital executive level then what we hear at an interdisciplinary meeting level. And I think one size does not fit all. I think one of the things we realized is that you have to take all that information and communication and adapt it to your local context. I think that's been very challenging ... (P10)*<br>*I really struggled with the communications. It would have been good to have a dedicated staff member in the ICU that was doing nothing more than channelling and filtering and then sending out the information that my staff require. The amount of time that we all spent looking at the same emails, trying to figure out what was new, what was relevant, what was important ... it would have been really good to have a dedicated staff member doing that ... integrating information across all of those clinical groups at the coalface that was almost a full-time job for an army of people ... that was done by clinicians. (P23)*<br>*If we just had one source of truth, I think it would be less confusing for people. (P10)*<br>*There were issues with the use of different types of masks — P2, N95, normal masks. And because there's different information coming from the Ministry, different information coming from the CEC [Clinical Excellence Commission] that was always lagging behind ... we almost felt that we were several steps ahead of the CEC, because CEC has to come up with a policy based on consensus ... Yes, there was confusion, come to think of it. (P31)* |
| d. Provide timely directives developed through transparent, credible and accountable measures | *I don't know where they got that evidence from. And, you know, that wasn't communicated to us. A lot of these policies were ... instigated overnight. And we just had to ... implement them, with little or no consultation. (P22)*<br>*Should have had some open forums or Skype forums where people could ask questions, because, while I knew what was going on, and the medical and ED knew, a lot of other people not in COVID wards didn't have a good grasp of what was going on. (P15)* |
|  | *So ... we called for hospital Q&A sessions ... they have an ID consultant in there ... and then at the end, it's [open for] questions. Just to assure the staff that this is happening, this is how we're going to deal with it. We're going to make sure that you guys are protected as a health care provider. (P32)* |
| **Theme 3. Plan for future crises** |  |
| a. Retain skilled multidisciplinary emergency taskforce | *There was a lot of confusion [not having enough staff] initially. A lot of anxiety and it slowed down workflow a lot. (P07)* |
| b. Implement written emergency preparedness plan with designated spaces and workforce | *I think preparedness is probably the biggest challenge. You never know what's around the corner. But to have contingencies in place, and to have a strategy that is not necessarily a tailored strategy, but it's a strategy that can act as a formula for a ready-to-go. (P13)*<br>*And when you have an evolving environment and COVID as well, it creates a lot of extra stress and uncertainty. When staff come to work not knowing whether maybe in two weeks' time we might move from this place and move to somewhere else again. (P09)* |
| c. Determine criteria and plan for escalation/de-escalation of clinical operations | *[Staff] have been rundown and then what we're going to do is, as soon as things ease, we're going to ramp surgery back up, which I felt was a bit of an issue. ... We could have stepped surgery back up in a clean, more controlled way. ... I think the most frustrating thing is ... there was no formal process of escalation. (P02)*<br>*We could have done [some preparation] prior to this pandemic, to some degree of how we would shut down services in a staged approach, where the priorities for staffing would be, activating flexible work plans, all that sort of thing. (P13)* |
| d. Deploy a multidisciplinary central control committee and a chain of command to facilitate communication, dissemination of information, and direct the review/development/update of guidelines, minimise the overflow of information and duplication of processes, and support guidelines appropriate to facilities' unique operations | *I still don't know what the hierarchy is, I report to all these different people. (P01)*<br>*There is a risk assessment, what level we're on at the moment ... at one stage, there's four levels to the risk assessment risk matrix that was developed here at [Hospital 2]. And then the CEC brought out a risk matrix with only three levels, and they didn't align. So we were often sitting on that line between green and amber. (P03)*<br>*Investigations are generally delayed ... It was such an ordeal to clean all the machines that would investigate them, certain services wouldn't even investigate the patients until they were [COVID] negative. And so, a lot of the time ... they weren't being investigated properly, they weren't being examined properly* |
|  | *... They go to fuel inpatient stay times ... other investigations ... unnecessary treatment ... huge follow-on effects. ... You just go for days ... until the swab came back negative ... definitely delayed their scans by 12 to 24 hours depending on the swab time. (P07)* |
| **Theme 4. Promote corporate and clinical agility** |  |
| a. Identify and protect vulnerable staff and patients to provide options, e.g. leave, redeployment, social support | *There are actually a lot of vulnerable healthcare workers around everywhere. For example, in my department, either older age, or some chronic disease, but they still work in the front line. I think they need to know, because of their risk, there's still a system in place for them. For example, if we need to relocate them to work in a lower-risk environment... (P09)* |
| b. Utilise alternatives to traditional team-based care to deliver quality care | *Their [Registrars'] regular training practices, their training clinics, their procedural lists were all thrown out the door with the ward-based system. I think one of the problems that we faced was that we felt that this ward-based system was thrust upon us. And even though there was limited consultation, I don't think there was any major consideration of the significant disadvantages of this system. ... It was a real black and white, all or nothing approach where we transition to immediate ward-based care or normal team-based care. ... We probably need to have a better consideration of how to stage our transition ... without going from one to the other extreme immediately. (P30)*<br>*"It [ward-based care] decreased the movement, but also made the patients in the ward a doctor's responsibility. So it led to earlier detection of problems and greater communication between the nursing staff and the medical staff, who knew what their patterns of communications were. So the doctors were more immediately available." (P06)* |
| c. Continue remote work culture that supports work-life balance (e.g. remote meetings and education where suitable) | *If we run a big departmental meeting, we can also invite other specialities to join and they just log in. That's it. You know, attendance is amazing, CPD [continuing professional development] will be amazing for 2020. ... The attendance at our departmental meetings is quite high because it's all Zoom ... virtual meetings are easy to log on to. So, again, it's easier to invite other specialities. (P21)* |
| d. Apply technologies such as telehealth to enhance patient care where applicable | *A lot of patients prefer ... telehealth because it's a big deal for some of our more frail or less mobile patients to get to the hospital, find parking, wait for their outpatient appointment, pick up their script. (P29)*<br>*It cost money to give them VPN [virtual private network] access, but we just went 'You know what, if you need to be able to see your patient records from home to be able to do your job and keep yourself safe [that works]'. (P29)* |

**Figure 1:**
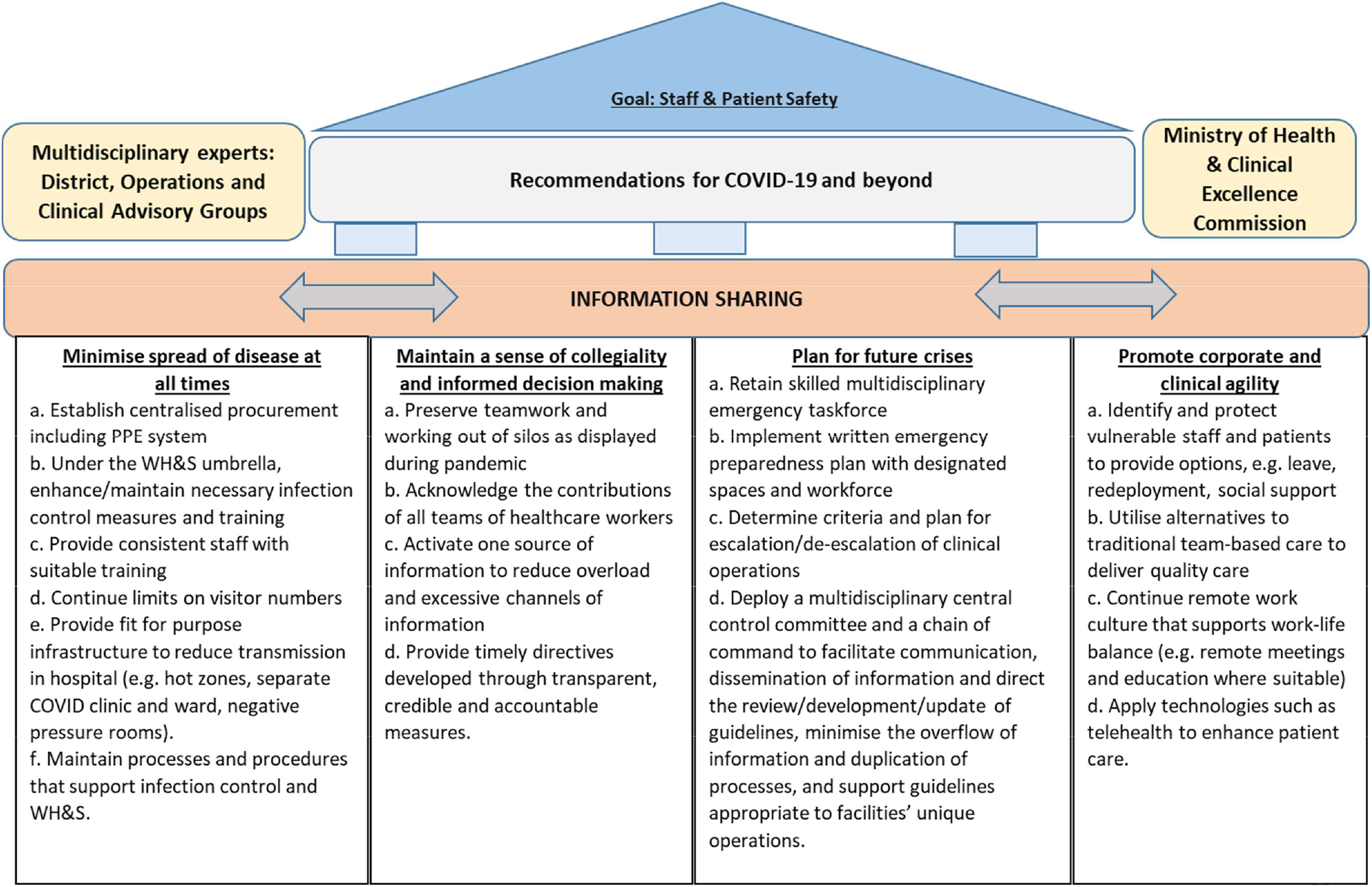
COVID-19 – Insights gained and recommendations for change.

(Fig 1: COVID-19 – Insights gained and recommendations for change)

### Theme 1. Minimise the spread of disease at all times

Combatting the COVID-19 pandemic requires caring for patients who contract the virus, while adopting infection control policies and procedures to protect the healthcare professionals who are enabling and administering care.

1(a) At the centre of staff concerns was the potential unavailability of PPE and uncertainties about the appropriate type and use of masks.

> *The contention around the mask … seemed to cause a lot of anxiety. (P11)*

The introduction of a centralised and managed PPE stock addressed multiple problems including staff stockpiling on wards, expired stock and escalating demands on the clinical product personnel to meet individual needs.

> *…and I think that [centralisation] worked well. You just made sure that you got what you needed for the week. (P14)*

1(b) The pandemic resulted in some participants advocating for aspects of infection control to be accommodated alongside Work Health & Safety (WH&S) practices that would keep staff and their patients safe year-round.

> *When it comes to keeping staff safe, you need to have a WH&S mindset, not an infection control mindset … Infection control relies on behaviours … WHS has a much more precautionary approach; it develops systems that, in my view, stop people from making mistakes. (P06)*

Conscious of the importance of PPE, staff demands for training/refresher sessions in donning and doffing created heavy workloads for nurse educators and led to innovations to address demands. Personnel were needed to organise the extensive implementation, including venues and schedules.

> *Part of infection control is education … [through] Grand Rounds [also]… on-the-spot PPE training on the ward. Anyone new coming on had to have a competency [assessment] done, … a lot of education [occurred] with individual departments, like radiology, the cleaners, ICU … (P36)*

1(c) Equipping the COVID ward with a permanent nurse unit manager and staff eliminated the daily task of training nurses from a temporary pool, and reduced time pressures on clinicians as well as the anxiety of patients whose questions previously went unanswered at best. Permanent staffing fostered teamwork and increased competency, valued in a highly stressful environment.

> *And so, you train someone up, and they’ll be gone the next day, you train someone else up, and they’ll be gone the next day … The patients they were helping would ask them questions, and they wouldn’t know the answers. Sometimes they just make answers up. And that’s when the shortcuts get made* … *(P01)*

1(d) The social isolation of patients who were already experiencing limited physical contact was distressing for all involved. Staff described the moral dilemma as they struggled to abide by the ‘no visitor’ policy, particularly for patients who were at the end of their life.

Acknowledging the unnatural caring situation, equipment and technology facilitated patient contact with family and loved ones. Staff struggled with, but appreciated, the need to limit visitor numbers.

> *Things like reducing visitor [numbers] and not allowing visitors … [entailed] big negotiation in the COVID ward. We were very firm about a lot of that stuff, no visitors, minimal people. (P03)*

1(e) Suitable physical infrastructure necessary for isolation such as reconstructing physical space to establish ‘hot zones’ for patients undergoing COVID testing was lacking initially. Challenges staff experienced in attempting to provide usual care included concerns about airborne transmission, the sanitising of equipment and staff’s restricted capacity while donned in full PPE.

> *It’s important that there are certain designated isolated areas for the different wards [for] patients who develop any symptoms which might be suspicious … cleaning equipment for the scanners if they need imaging, and the right workflows to ensure … imaging if needed. Having the right spaces to keep the patient so that clinicians and nursing staff feel comfortable treating them and reviewing them, examining them and providing plans and updates in person. (P07)*

1 (f) Throughout the pandemic, minimising the risk of airborne transmission has been a priority. Risky procedures such as the use of nebulisers and non-invasive ventilation were stopped, and alternative or new procedures were introduced.

> *Simple things, like the use of PPE, avoiding nebulisers, the process of intubating people, …non-invasive ventilation [NIV] have changed forever. The processes of cohorting people and risk stratifying at triage, that’s something that’s going to change forever. A lot of practice has changed … those changes will be permanent. (P31)*

### Theme 2. Maintain a sense of collegiality and promote informed decision-making

Effective management enabling a quick pace of change across the facility, most consequential during the initial uncertainties such as virus-host interaction and length of period of transmissibility, was dependent on the leadership of and collaboration among staff and fit-for-purpose communication methods, channels and communicators. The majority of participants, including the Heads of Departments, spoke passionately about frequent and productive inter-department and department-executive interactions that permitted ‘on-tap’ problem solving for the continuation of a range of routine services employing innovative approaches. A critical facilitator was the creation of three hierarchical committees, one of which was the Clinical Expert Advisory Group (CEAG), which met as frequently as the situation required, daily or more often, and continues to meet more than a year and a half on from its inception.

2(a) Leadership and the teamwork it supported were essential for timely decision-making and the continuation of health care delivery during the pandemic.

> *The strength of our response has been the camaraderie and goodwill between clinical services. …teams have really pulled together wonderfully well. Never had any pushback about COVID and who’s taking what responsibility. I think everyone stepped up remarkably. (P10)*

Leadership and strong collaboration evident in the CEAG meetings were widely acknowledged and enabled timely solutions and directives to address critical issues.

> *I think maintaining some of those meetings, where there is a broad input from clinicians to executives in an open format, would be helpful. (P06)*

2(b) The pandemic placed urgent and extensive demands on staff, the majority of whom stepped up to the challenges. A recurring theme was unscheduled work hours with demanding workloads. Unsurprising were calls for acknowledgement of outstanding contributions of staff.

> *I feel like there’s a lot of unsung heroes. (P12)*

2(c) Information overload from numerous local and international sources, and traditional and social media, often exacerbated staff anxiety and created an onerous task for those responsible for policies and guidelines. Conflicting information, for example about the appropriate type of and circumstances for wearing a mask, was challenging for most.

> *We had WHO guidance, we had advice from the federal government, from the AHPPC [Australian Health Protection Principal Committee], from the Ministry [of Health], who had advice from the Community of Practice. So much advice, so many documents being drafted and redrafted and redrafted. (P18)*

Managers had a burdensome task to justify the reason for inconsistent policies across the state and among Local Health Districts (LHDs). The consequence was heightened fears that inhibited optimum health care and, in some circumstances, led to deficient patient care.

> *I think we were really challenged because the different LHDs did things differently. And some people who worked both, say, here and the LHD2, would come back from LHD2 going, ‘Oh, they’re doing this?’ And I [would say] ‘Well, you know, we are very clear that we will do…*.*’ I think that was really challenging, having variation between LHDs. So I think at a state level that should be done differently. (P29)*

2(d) Transparency, credibility and accountability of timely directives were noticeably absent. The initial lack of identified channel(s) of trusted information resulted in misinformation, anxiety and inconsistent practices. Participants voiced concern about operational decision-making that sometimes lacked input from staff at the coalface.

> *One of the things that was potentially lacking was transparency with what was happening at the state level, or within the Ministry of Health level, that appeared to be almost like a black box. (P06)*

Missing from state directives was an appreciation that health facilities were not equal pandemic responders and therefore directives needed to be specific and detailed..

> *They did have an intensivist from [Hospital1], who was part of the advisory group … But [Hospital1] really didn’t have much direct input into the day-to-day management of COVID. It was really [Hospital2] managing that…there was a lack of input of [Hospital2] within the Ministry’s decision-making, representing essential intensive care. (P06)*

Timely dissemination of information via the Chief Executive’s Broadcasts to all HCWs gave credibility to the rapidly changing workplace requirements. Open forums such as Q&As and Grand Rounds were valued because they allowed staff to seek answers and have further clarity. Some departments also employed additional mechanisms to keep staff informed.

> *I think the communication was a way of relieving staff anxiety …the Chief Executive’s daily broadcast was quite helpful … Grand Rounds initially was very effective. The Infectious Diseases team was so articulate, and they gave such measured responses, and gave people the information that they needed. (P03)*

### Theme 3: Plan for future crises

The ambiguity surrounding appropriate procedures and directives for the provision of care in a safe environment fuelled feelings of stress and anxiety. Participants described the benefits of a multidisciplinary team armed with the expertise, experience and resources capable of providing an immediate response and ongoing governance and capacity for the wide-ranging components integral to a comprehensive pandemic response.

3(a) COVID-19 demanded an urgent review of usual health care activities. Delays were counterproductive to efforts to keep staff engaged. A taskforce that could be activated at a moment’s notice would minimise delays, staff anxiety and the accompanying risks to staff and patient well-being.

> *Having an emergency taskforce at the hospital where, say, if anything like this happened—Ebola, some sort of pandemic, a fire—you have ten designated nurses that you would pull out of different departments … a mobilised taskforce with consistent nurses* … *all ready from different wards that had already been designated. … And these are the [taskforce] doctors. (P01)*

3(b) A pandemic plan detailing components for a comprehensive response would minimise delays and uncertainties. Details would support a proactive response and identify designated spaces appropriate for isolating and caring for suspected or infected patients.

> *We should have had a pandemic plan. We have for influenza; we’ve had these plans … at an executive level probably sitting on a shelf somewhere…. But it’s not just about having a document as to how you do these things … we’ve learned …we were doing things very reactively. But now, part of our mandate with the biocontainment centre is that we’re going to be thinking very proactively about pandemic planning, and at every level… (P10)*

3(c) The criteria and a plan for modifications to clinical operations, including escalation and de-escalation of procedures, would enable staff to be prepared and act in unison.

> *We probably need to have a better consideration of how to stage our transition from a team-based or ward-based system without going from one to the other extreme immediately. (P30)*

3(d) Clarity around roles, responsibilities and lines of reporting were blurred or absent during the height of the pandemic, deleterious to patient care and staff well-being. Long hours were spent developing solutions for working at the coalface that were superseded by less fit-for-purpose directives from higher levels of the health sector. A multidisciplinary central control committee could efficiently create expert groups with the authority and responsibility to action urgent and ongoing change.

> *I think there has been a very strong case for us to have some sort of strong national, multidisciplinary communicable disease centre, where you incorporate all the leaders from these various groups into one entity that could then share their information and make sure that all the information that they’re sharing with their groups is consistent. (P10)*

### Theme 4: Promote corporate and clinical agility

Staff and patient safety and well-being are paramount concerns for any health system. Circumstances during the pandemic fashioned unique responses to the provision of health care and staff’s ways of working.

4(a) Individual staff/patient characteristics and needs were foremost in the minds of those determining the day-to-day operations in the facility. Staff’s age, personal circumstances and physical and mental status were key considerations in determining their vulnerability, and simultaneously, their capacity to contribute to the facility’s pandemic response. Similarly, confidence in staff enabled flexibility in caring and advocating for patients.

> *The majority of the time [visiting rules] were followed. There were a few exceptions, though, where families did try and do their own thing…. The hospital staff did the best they could under the circumstances…. Sometimes it’s not possible though, especially if you have someone who’s very sick or dying … that’s the hardest bit. (P16)*

4(b) Perspectives on adopting a ward-based model of care, while diverse, were predominantly positive. Concerns were voiced about staff in training achieving required competencies and the immediacy with which the model was introduced. Conversely, the medical staff were more easily accessible and shifts that accompanied the ward-based care minimised un-rostered and overtime schedules.

> *Changes from the medical staff model to your ward-based group, they should try and hang on to that, but that’s going to take some investment of resources. … That really made a difference. … But that’s completely changing the delivery of medical care…. (P20)*

4(c) Technology offered the organisation an avenue to sustain aspects of workplace requirements, as staff could remotely attend meetings, engage in educational sessions, review medical notes and communicate with patients. The remote work culture was welcomed by most, despite its sudden and widespread application.

> *It was a significant cultural shift for the entire organisation to realise that you don’t need to physically be in the building to be able to contribute. There’s a role for physically walking around and seeing the lie of the land. But there are also certain advantages, not spending hours travelling to and from an office … you can be equally productive in this sort of setting. (P06)*

4(d) Technologies such as telehealth to enhance patient care were welcomed by most clinicians, with some advocating for continued or greater use. Telehealth supported self-isolation, reducing risk of exposure and spread of COVID-19. Senior clinical staff touted the value of a permanent virtual ED.

> *I’ve found telehealth really useful. I do everything by telehealth now. (P17)*
>
> *I think telehealth could have been done better, to be honest. I think we could have had [a] virtual emergency department for patients who were suffering conditions where they needed to see a doctor … to be in an emergency department, but they couldn’t because of COVID. … And then they could have had a more planned approach. (P34)*

For certain conditions, such as those requiring a physical examination or patient observation, telehealth was deemed unsuitable.

> *We decided that there were very few visits that we could cut out, or that we could move to video. …we needed to keep doing most of these visits as face to face visits, because to do otherwise was going to potentially make things unsafe. (P38)*

## Discussion

Australia’s consistent easing of restrictions from late 2020 signalled the opportune time to capture the lived experiences and perspectives of staff that could inform improvements to quality health care now and into the future. We aimed to identify successes to inform facets of health care that should be maintained, and at the same time, document into corporate memory aspects that should be modified if not entirely avoided. Our findings focus on the well-being of HCWs and patients, and aspects of governance and management of a tertiary hospital.

### Considerations for ongoing and future provision of health care Minimise risk of infection

Health care organisations around the world are reassessing how to safely provide essential care for patients in times of crisis.[22] The pandemic elevated the importance of staff safety and prevention of transmission to the community comparable to patient safety.[23, 24] Heightened anxiety – stemming from the unknowns of the virus,[3] inconsistent guidelines, and inadequate PPE stocks and directives – was not unique to Australia.[25] Sudden and extensive demands for training stretched the resources of Infection Control and nurse educators. Establishment of a centralised managed stock enabled transparency and ready access, thereby reducing individual hoarding and stockpiling.

The utmost priority given to infection control positively affected WH&S aspects of the healthcare system. Posited as the way forward, infection control practices could be effectively incorporated into routine activities if perceived as a WH&S issue, a concept previously reported.[26] One such example, initiated by the ED, saw staff assigned designated workstations and computers and required to declutter and regularly clean surfaces to reduce contamination and sources of infection. Restricting visits, employed as an early infection control strategy, caused moral injury,[27] particularly for staff unable to apply their professional judgement. Guidelines enabling patients to safely receive visitors[28] reflect an appreciation for the benefits of visitors to patients[29] as well as the need for regulation.[30, 31]

Efforts directed towards procedural changes and staffing may have relevance to non-pandemic operations.[32] Staff agility enabled the adoption of rapid changes, some temporary as was an innovative process for intubating patients, and others long-term such as avoidance of nebulisers, cohorting patients suffering from infectious diseases and use of suitable physical infrastructure. Work is already underway to review regulations and standards on hospital buildings and the guidelines that govern their operations.[33, 34] Minimising the contact between infected and non-infected patients and staff using ‘shelter hospitals’[35] would enable non-pandemic operations to continue, thereby minimising the de-escalation of services.

Early designation of a separate testing clinic and ward, the latter equipped with permanent staff and a Nurse Unit Manager (NUM), would support operations, staff and patient safety, and work demands of assigned medical practitioners. Pool staff increased the likelihood of shortcuts of various procedures, unauthorised staff providing directives, PPE breaches, and anxiety and unabated fear among both poorly informed staff and patients.

### Maintain a sense of collegiality and informed decision-making

The general motivation of staff to be involved early in the pandemic might have stemmed from feelings of obligation in the presence of facilitators such as the provision of accommodation, and at a local level, transparent information sharing and a sense of inclusion in decision-making,[36, 37] which would have alleviated feelings of distress that were common among HCWs.[38] The collaboration participants experienced, desirous across occupational and professional groups,[39] was underpinned by multipronged communication strategies between staff at the coalface and the leadership groups, and was particularly valuable in the periods of rapid change, as noted elsewhere.[40] Recounts of everyone having the same agenda were as common as those of un-rostered hours worked – both warranting recognition.

### Plan for future crises

Despite SARS-CoV-2 being the third coronavirus to emerge in the past 20 years and experts previous warning of impending deadly epi/pandemics,[41-43] the world was unprepared for COVID-19. In addition to ‘a detailed operational blueprint’[43] for an effective response, participants expressed the importance of the retention of a skilled multidisciplinary emergency taskforce and, for sustained analytical and operational capacities, a multidisciplinary central control group. A taskforce would prove invaluable particularly in situations when there is an extended period before the infectious agent is identified such as those presented by COVID-19, compared to the SARS outbreak when researchers quickly identified the infectious agent as SARS-CoV-1.

Up-to-date information, education and training are not only prerequisites for quality patient care but are important for reducing HCWs’ risk of psychological distress.[44] Integrated in the control centre, designated individuals in core response areas such as ICU[31] could establish information channels on the current status of operations that would strengthen collaboration and coordination across diverse but relevant groups. Hand-in-hand with information sharing and having what has been referred to as a *transparent strategy*,[45] is acknowledging uncertainty[46] to abate confusion and panic.

An effective control centre would: engage frontline staff and experts across disciplines and levels of governance[47] in decision-making and the development of evidence-based policies and protocols,[35] avoiding duplication of processes; prioritise infection control, protection and safety measures ahead of knowledge of disease transmission;[38] create central stocking and stockpiling of equipment to avoid resource strain;[48] and deploy standardised frameworks to keep all staff abreast of updated protocols, guidelines and policies. Acknowledged as a key component of a preparedness plan is a scalable emergency system capable of responding to a surge in demand for resources and patient care.[17, 49]

### Promoting corporate and clinical agility

Participants extoled the agility of staff as they reflected on efforts to achieve essential elements for a responsive health care environment that provides quality patient care while simultaneously mitigating the spread of the virus amid rapid and frequent change. Similarly, leaders acted swiftly to mitigate risk, introducing a ward-based model of care aimed at minimising the movement and interacting of staff at work, and offering assistance to staff identified as vulnerable – the latter a known effective measure.[50] Risk management included staff taking leave, working remotely and being redeployed. Working remotely, a practice previously considered impractical,[51] especially in health care related jobs, became a reality for many in the pandemic.[52] However, still lacking are appropriate guidelines for individual sectors[51] and assessment of its benefits.

Telemedicine proved effective in filling some health care gaps, enabling healthcare services to infected and non-infected people during the pandemic.[53] In our study clinicians had mixed reactions to its sustainability, citing situations when a traditional physical examination is necessary. Nonetheless, most appreciated that remote working has the potential to be cost-effective, extend access to specialist care,[54] and increase productivity and job performance.[55, 56]

### Limitations

The present study focused on one facility that is a designated COVID-19 facility in Australia, potentially limiting its generalisability. Our purposive sampling however, captured key informants who were representative of crucial sectors of the pandemic response. They were strategically placed to identify successes and gaps in the provision of health care, and to make considered recommendations reflecting both a bird’s-eye view and coalface experiences. Despite never reaching the dire situation experienced elsewhere, it can be argued that the four themes hold worldwide applicability – with a number of their sub-components identified as important considerations in studies from Europe,[35] Canada,[49] and USA[15] – and contain generic concepts with international relevance. Our methods employing Morrow’s et al.[20] modification of Colaizzi’s phenomenological approach were strictly adhered to and re-analysis was undertaken, culminating in results based on four researchers to minimise researcher bias.

## Conclusion

This study presents, to our knowledge, the first report of lived experiences and recommendations from clinical and non-clinical senior healthcare professionals in Australia. Their observations and recommendations should inform decision-makers tasked with mobilising an efficacious approach to the next health crisis and, in the interim, aid the governance of a more robust workforce to effect high quality patient care in a safe environment. Admittedly, the initial challenge rests with leaders who must agree to prioritise fit-for-purpose systems and structures as part of crisis preparedness while simultaneously tackling inexorable current demands.

## Data Availability

Data produced in the present study are not available as participants will be potentially identifiable.

## Acknowledgments

The authors would like to thank Nicole Tolhurst and Elizabeth White for their guidance in the study’s approach, all participants for their time and considered responses, Peter McCaul for reviewing and providing useful feedback on manuscript drafts, and Leendert Moerkerken who identified and facilitated the transcription process.

## Notes

### Competing Interest Statement

The authors have declared no competing interest.

### Funding Statement

This study did not receive any funding

### Author Declarations

Western Sydney Local Health District Ethics Committee. Ethics approval granted (2020/ETH01674)

